# Lethal phenotypes in Mendelian disorders

**DOI:** 10.1101/2024.01.12.24301168

**Authors:** Pilar Cacheiro, Samantha Lawson, Ignatia B. Van den Veyver, Gabriel Marengo, David Zocche, Stephen A. Murray, Michael Duyzend, Peter N. Robinson, Damian Smedley

## Abstract

Essential genes are those whose function is required for cell proliferation and/or organism survival. A gene’s intolerance to loss-of-function can be allocated within a spectrum, as opposed to being considered a binary feature, since this function might be essential at different stages of development, genetic backgrounds or other contexts. Existing resources that collect and characterise the essentiality status of genes are based on either proliferation assessment in human cell lines, embryonic and postnatal viability evaluation in different model organisms, and gene metrics such as intolerance to variation scores derived from human population sequencing studies. There are also several repositories available that document phenotypic annotations for rare disorders in humans such as the Online Mendelian Inheritance in Man (OMIM) and the Human Phenotype Ontology (HPO) knowledgebases. This raises the prospect of being able to use clinical data, including lethality as the most severe phenotypic manifestation, to further our characterisation of gene essentiality. Here we queried OMIM for terms related to lethality and classified all Mendelian genes into categories, according to the earliest age of death recorded for the associated disorders, from prenatal death to no reports of premature death. To showcase this curated catalogue of human essential genes, we developed the Lethal Phenotypes Portal (https://lethalphenotypes.research.its.qmul.ac.uk), where we also explore the relationships between these lethality categories, constraint metrics and viability in cell lines and mouse. Further analysis of the genes in these categories reveals differences in the mode of inheritance of the associated disorders, physiological systems affected and disease class. We highlight how the phenotypic similarity between genes in the same lethality category combined with gene family/group information can be used for novel disease gene discovery. Finally, we explore the overlaps and discrepancies between the lethal phenotypes observed in mouse and human and discuss potential explanations that include differences in transcriptional regulation, functional compensation and molecular disease mechanisms. We anticipate that this resource will aid clinicians in the diagnosis of early lethal conditions and assist researchers in investigating the properties that make these genes essential for human development.

## Introduction

### Defining essentiality and sources of evidence

Essential genes are defined as those required for growth, proliferation, and survival of a cell or an organism. The resulting set of genes being labelled as essential will vary significantly depending on the level of organisation being considered (cell, organ, whole organism), the species, or the exact definition/ thresholds used, e.g., a quantitative gene effect score in different cell lineages. Cellular essential genes are those required for cell proliferation while lethal genes in the mouse are defined by the International Mouse Phenotyping Consortium (IMPC) as those where homozygous knockouts die during embryonic development or soon after birth, during the pre-weaning stage ^1^. Similarly, in humans, lethality can be investigated prenatally, which provides information on the essential nature of genes for early organism development ^2,3^. From an evolutionary perspective, lethal genes could be defined as those required for growth to a fertile adult, i.e. affected individuals die before reproductive age in the absence of treatment, or even genes leading to physical and intellectual phenotypes that impede reproductive success ^4^. The complete loss-of-function (LoF) of a gene may lead to a wide spectrum of phenotypic abnormalities ranging from: clinical infertility due to embryonic loss at the earliest stages of development, i.e. embryonic lethality before a pregnancy is clinically recognised; prenatal lethality; multiple neurodevelopmental, metabolic and skeletal disorders that may result in neonatal, infant or childhood death; disorders with later age of onset associated with premature death; abnormal phenotypes that may not impact life expectancy; or even no detectable clinical phenotypes.

Moreover, gene essentiality is not an absolute or binary trait. Even when using exactly the same definition, and considering the same organism and organisational level, gene essentiality may be context or tissue specific (gene is essential only in specific cell types) or genetic background specific, e.g. discrepancies in mouse viability have been found for up to 10% of genes when knocked out ^5^. In the presence of the same genetic background and null allele, lethality can manifest with incomplete penetrance ^1^. Another aspect involves essentiality and allelic requirement. Traditionally, essentiality has been evaluated in homozygous knockouts, however there are haplo-essential genes that cannot tolerate a LoF mutation in one or both alleles ^6,7^. This set of genes is not as well characterised in mouse knockouts, and the comprehensive approach of the IMPC presents an opportunity to explore this further.

The current evidence on essential genes in humans comes from various sources providing insights into the phenotypic impact on a gene’s LoF at distinct levels, as recently reviewed ^8^ and illustrated in **Figure 1**.

i) Genes essential for cell proliferation, identified primarily through RNA interference and CRISPR-Cas9 disruption in human cancer cell lines ^9^. More screens are now being performed in human pluripotent stem cells (hPSCs) ^10,11^. Defining a ‘core’ set of essential genes is not straightforward and several thresholds and approaches have been suggested ^12^.
ii) Intolerance to variation metrics derived from large scale human population sequencing programmes ^6^. Notably, these scores provide a measure of how intolerant to (heterozygous) LoF a gene is and how likely it is to underlie single gene disorders, but not on the nature or severity of the phenotype. Recent metrics are being estimated from different datasets and incorporate machine learning approaches that integrate additional gene features ^13,14^. Again, recommended thresholds can be used to identify highly constrained genes. The presence of homozygous LoF variants ^15^ or the deficit of homozygosity among protein-altering variants in the general population ^16^ are other examples on how information from large sequencing studies can be used to understand gene constraint and potential phenotypic impact.
iii) Resources compiling clinical reports on single-gene disorders that enable users to conduct queries based on phenotypic criteria, including the Online Mendelian Inheritance in Man ^17^ (OMIM), the Human Phenotype Ontology ^18^ (HPO) and the Monarch Initiative ^19^ repositories. In this case, the essential nature of a gene is defined by records of early death.
iv) Human orthologues of essential genes in different unicellular and multicellular organisms, especially genes that are essential for mammalian organism development ^20,21^. In the mouse, this information can be obtained through different viability screens that assess the viability of homozygous knockouts during embryonic development and the pre-weaning stage ^7,22^.

**Figure 1.**
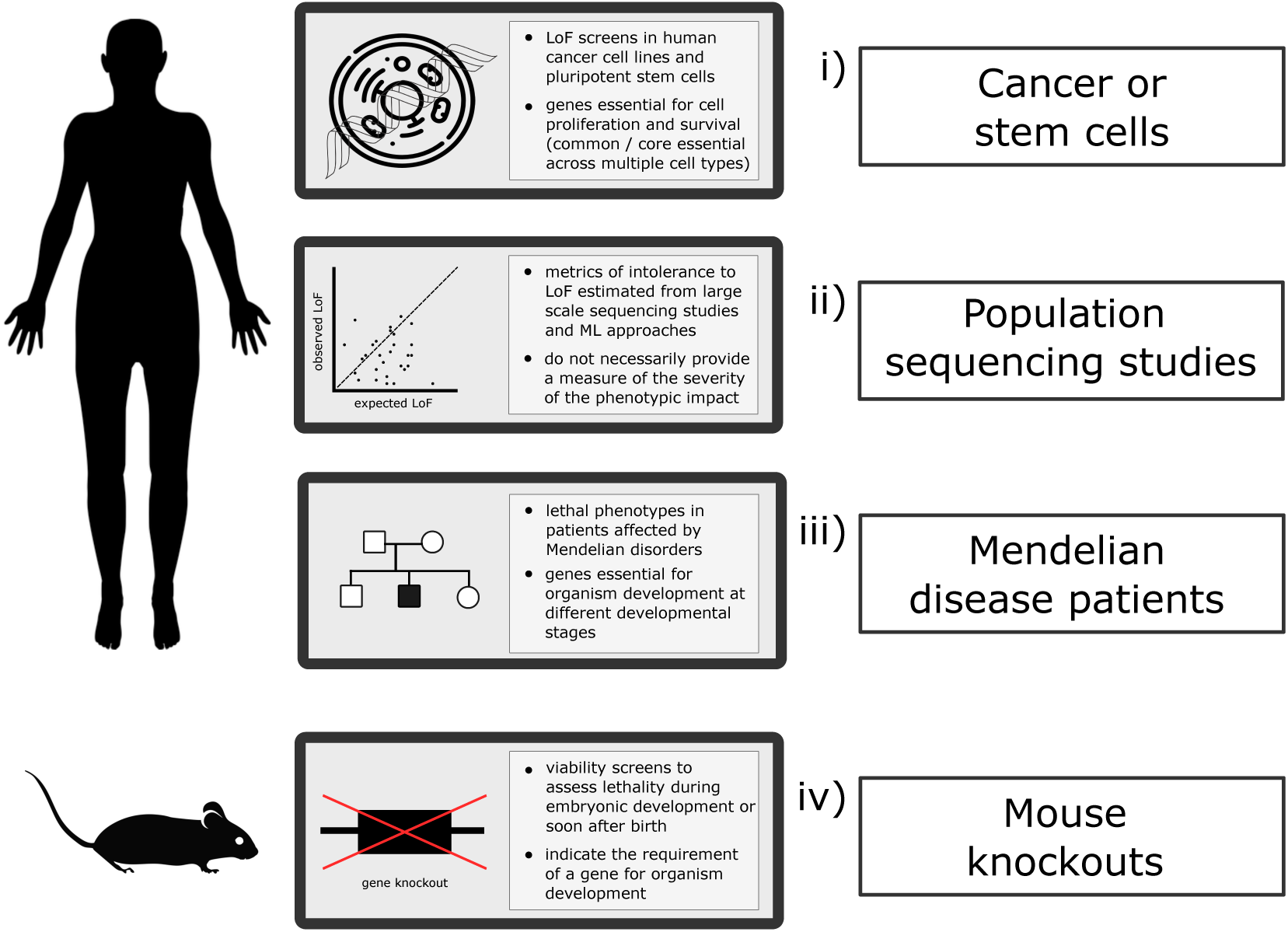
Current sources of gene intolerance to LoF variation/essentiality and lethal phenotypes. The phenotypic impact of a gene’s LoF can be assessed in humans at the cellular level, through cell proliferation assays; observed versus expected variation inferred from large scale population sequencing data and evaluation of lethal phenotypes in patients affected by Mendelian conditions. In the mouse, different viability assessment screens allow the identification of homozygous knockouts with preweaning lethal phenotypes. LoF: loss-of-function; ML: machine learning.

### How can knowledge of gene essentiality inform human disease studies?

Genome sequencing has revolutionised the molecular diagnosis and clinical management of patients affected with rare disorders, yielding earlier and better personalised therapies and providing the opportunity for reproductive counselling. Options available for patients with an autosomal dominant (AD) condition or carriers of autosomal recessive (AR) disorders include preimplantation genetic testing for monogenic disorders (PGT M), prenatal and newborn testing. Despite these successes, many patients still remain undiagnosed as a clearly pathogenic variant in a gene with proven involvement in the condition is not discovered, e.g., some 75% of cases in the 100,000 Genomes Project remain unsolved ^23^. Around 20-25% of the protein coding genome is recognised to be associated with Mendelian disorders ^17^. Recent estimates suggest the final number of Mendelian disease genes will be 1.5-3 times higher ^24^. Among the strategies to identify novel Mendelian genes and diagnose more patients, in particular those presenting with prenatal and early neonatal phenotypes, mouse knockout databases constitute a source of candidate genes linking phenotypic outcomes, including prenatal lethality, to homozygous and heterozygous LoF ^25^.

Mouse knockout lines with viability information are now available through the IMPC resource for up to one third of the protein coding genome, and Mendelian disease genes are significantly enriched for lethality in the mouse ^1,5,26^. This observation is consistent across different modes of inheritance and rare disease categories as established by PanelApp, an open knowledgebase of virtual gene panels related to human disorders ^27^. However, the proportion of lethal genes is not evenly distributed across disease classes. A higher number of affected tissues and physiological systems has been found in disorders associated with genes that are lethal in the mouse ^5,28,29^ (**Figure 2**).

**Figure 2.**
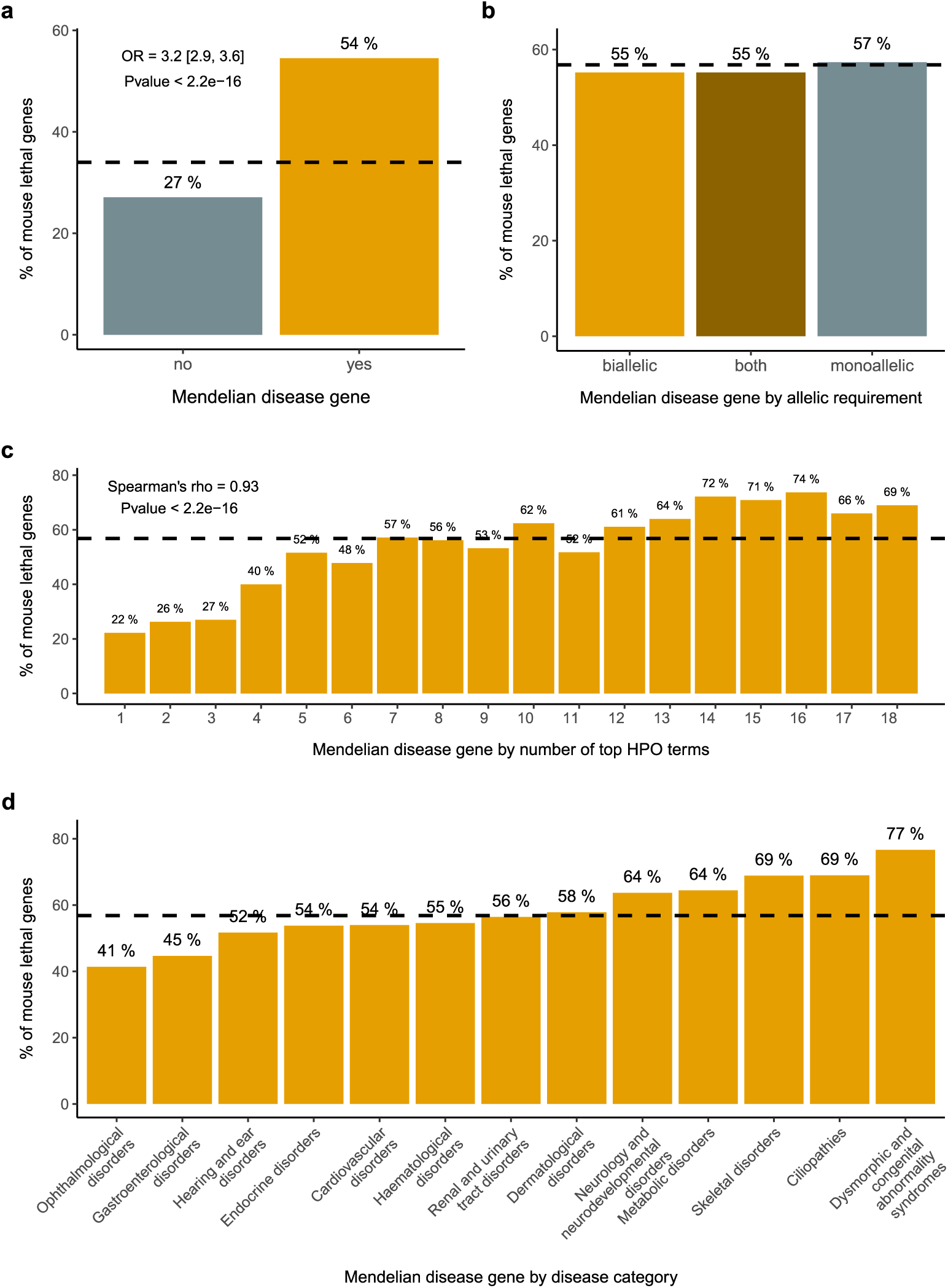
Association between Mendelian and mouse lethal genes a) Mendelian genes and mouse viability. The set of Mendelian genes is significantly enriched for mouse lethal genes (OR 3.2; Pvalue < 2.2e-16) **b) Mode of inheritance and mouse viability** No significant differences are observed when disease genes are categorised according to the associated allelic requirement **c) HPO terms and mouse viability** The percentage of mouse lethal genes among Mendelian disease genes is correlated with the number of physiological systems affected, as captured by the number of high level HPO terms (Spearman’s rank correlation rho = 0.93; Pvalue < 2.2e-16) **d) Disease categories and mouse viability** The percentage of mouse lethal genes is not uniform across disease categories. Mouse ethal genes from IMPC DR20.1 viability assessment (lethal + subviable); Mendelian disease genes, allelic requirement and disease category for those genes present in PanelApp with significant clinical evidence (green, diagnostic-grade); HPO top terms from gene-to-phenotypes Human Phenotype Ontology annotations. The dashed lines represent the baseline percentage of knockout lines with a one-to-one human orthologue that are lethal (lethal + subviable) (a) and the percentage of lethal (lethal + subviable) lines among Mendelian disease genes (b),(c),(d).

When we break down the set of mouse lethal genes into more granular categories, i.e. cellular lethal and developmental lethal genes ^5^, or early gestation, mid gestation and late gestation lethal genes ^30^, we observe that this enrichment is not uniform, and that the sets of developmental lethal or late gestation lethal genes are the ones more significantly enriched in human disease genes. This observation led to the hypothesis that the set of cellular or early gestation lethal genes may be associated with early embryonic lethality in humans, a phenotype not well captured in current gene-disease association databases and difficult to ascertain, as it will be explained in the next section.

Where human mutations in genes, that have been shown to be lethal in the mouse, result in a lack of prenatal lethality in patients, it could be that there is not a total loss of protein function, i.e. heterozygous LoF or hypomorphic variants that lead to reduced protein function and could manifest as postnatal phenotypes ^28^. Even when humans do not exhibit the extreme phenotype of lethality associated with mouse null alleles, the mouse embryonic manifestations and postnatal abnormalities in the early adult heterozygous knockout contribute to facilitating the interpretation of variants identified in humans with different molecular consequences, variable penetrance and/or expressivity and to understanding disease mechanisms ^31^. Overall, the set of genes that are lethal in the mouse and not currently associated with human disease constitutes a powerful source of genes potentially linked to severe, multisystemic, early onset disorders in humans, including those with prenatal and neonatal lethal phenotypes, e.g. families with a history of miscarriages followed by a child with a severe, early age of onset condition of suspected monogenic origin ^5,26,30,32^. Gene and variant prioritisation strategies leveraging this information have been successful in identifying novel neurodevelopmental disease genes ^33,34^. Similarly, together with other metrics of intolerance to LoF, information on mouse essential genes is a highly predictive feature in machine learning implementations to predict disease risk genes ^35^.

### The challenge of identifying lethal genes in humans: diagnosing lethal fetal disorders

Prenatal exome, and more recently, genome sequencing has been introduced to routine clinical care for at-risk pregnancies in which a genomic diagnosis would guide management of the foetus ^36^, and in the extreme case of prenatal death, to perform molecular genetic testing to determine the genetic cause of pregnancy loss or perinatal death ^37^. Similar to later onset phenotypes, an important proportion of pregnancy losses lack a molecular diagnosis. Microarray analysis of sporadic and recurrent pregnancy loss samples did not detect clinically significant chromosomal abnormalities in ∼ 42% of the samples, with pregnancy losses occurring during the earlier stages of gestion being more likely due to such genomic imbalances. In the case of stillbirths, the percentage of cases with non-chromosomal abnormalities goes up to 85-90% ^38^. Remarkably, stillbirths were recently found to be enriched for LoF variants in genes not (yet) associated with Mendelian disease, compared to undiagnosed patients with postnatal manifestations ^3^. All this evidence suggests that pregnancy loss in euploid pregnancies can often have a Mendelian or polygenic origin, indicative of the ‘essential’ nature of the implicated genes. Robbins et al. suggested that lethality occurring early in the pregnancy would involve genes related to fundamental cellular processes, while genes associated to developmental biological processes would be associated to death occurring at a later fetal developmental stage ^39^. The analysis of mouse knockout lethal genes confirms the association between the embryonic stage at which lethality occurs and the different biological processes ascribed to each corresponding genes ^5,30^. It is important to emphasize that while mouse embryonic stage refers to any developmental stage before birth, in humans there is an embryonic and a fetal stage.

Abnormal prenatal phenotypes may be limited to fetal life and are not thoroughly investigated or clinically characterised since the outcome consistently involves prenatal or perinatal lethality. However they can also manifest as a more severe form of a previously identified postnatal-stage phenotype, posing a challenge for molecular diagnosis during this developmental stage ^40^. Regarding pre/perinatal lethality, we anticipate identifying a set of genes in which this is the sole observable phenotype, as well as genes where early lethality is the most severe manifestation within a broader spectrum of postnatal phenotypes. Consequently, when investigating potentially pathogenic variants associated to prenatal death, one of three scenarios are possible: i) the gene is a known Mendelian gene and the lethal phenotype association has been previously described, ii) the gene is a known Mendelian gene with postnatal (and/or other prenatal) manifestations and the prenatal lethal phenotype constitutes an expansion of the phenotype, iii) the gene is not yet known to be associated to a single-gene disorder. Allelic presentation needs to be factored in, e.g., complete vs partial LoF, biallelic lethal vs monoallelic viable with other less severe postnatal manifestations ^41,42^. It is worth noting that most of the homozygous lethal mouse knockouts present a viable but abnormal phenotype in the heterozygous state (1497/1891, 79%; IMPC DR20.1). The actual percentage may be even higher as not all the lethal lines have completed the planned phenotyping screen for the corresponding heterozygous knockout, or if additional phenotypes were explored.

Many fetal demises are sporadic and a monogenic cause may not be suspected, therefore molecular genetic testing is either not performed or it does not provide a definite diagnosis. This makes it impossible to establish gene-phenotype associations for these cases ^42^. Previous evidence supports a model where highly intolerant to LoF variation genes are not known to be associated with recognisable human phenotypes due to the fact that the resulting phenotype is always early embryonic death^5,32,43^. In this scenario, monogenic forms of this extreme phenotype of lethality are likely underrepresented in current disease databases.

The lack of a fetal phenotype resource, equivalent to those assisting the molecular diagnosis of postnatal disorders, adds to the challenge, although efforts are being made in that direction, i.e. the creation of a Prenatal HPO working group as part of the efforts of the Fetal Sequencing Consortium ^44^ and the submission of new fetal phenotype-genotype associations to facilitate variant curation ^45^. Given the increasing number of prenatal and molecular autopsy sequencing studies, the number of prenatal and perinatal lethal phenotypes associated with known and novel Mendelian genes is expected to increase.

### The need for a catalogue of lethal phenotypes in humans

First, as described above, the direct evidence for human essential genes comes mainly from cell proliferation assays in human cancer cell lines. While these genes perform functions required for cell survival, i.e., ‘cellular lethal’ genes, there is evidence of a larger set of genes playing an essential role in developmental processes, i.e., ‘developmental lethal’ genes. This evidence, supported by postnatal and embryonic viability screens in mouse, suggests that the set of human cellular essential genes identified through the first approach will not necessarily capture those genes essential for human development beyond the earliest embryonic divisions ^5^.

Second, the number of molecular autopsies, sequencing studies of fetal structural anomalies, often severe and lethal, and those aimed at identifying genetic variants associated with pregnancy loss and perinatal death is increasing ^3,37,42,46,47^. However, potential pathogenic variants identified through these studies are not adequately collated and easy to query for subsequent diagnosis.

Some attempts have been made to try and identify these prenatal lethal phenotypes from the literature ^48^. One informatic toolkit developed by Dawes et al. retrieved data from different sources to generate a list of genes considered candidates to be associated to unexplained infertility and prenatal or infantile mortality ^26^. Information from this resource combined with mouse evidence and LoF variants documented in the gnomAD database has recently been used to generate a candidate set of genes related to human lethality and pregnancy loss and compute carrier rates of pathogenic/likely pathogenic variants in those genes ^49^. There are several resources for single-gene disorders that enable users to perform phenotypic queries, including OMIM and the HPO repositories. The available evidence documented for conditions characterised by prenatal and perinatal lethal phenotypes in humans is either scarce or difficult to extract from these knowledgebases. A previous study based on OMIM reports 624 genes with perinatal lethal phenotypes ^26^. The most recent HPO release (v2024-01-11) contains 457 genes and 514 disorders with an age of death annotation, likely an underestimation ^18^. The information currently available from OMIM on lethal phenotypes is not captured in a comprehensive manner, mainly being described in heterogeneous free text reports. Establishing the link with lethality is not always straightforward from the information available in this resource, e.g., ambiguous cause/age of death, reports of prenatal lethality of siblings/death of other family members with no molecular diagnosis and only possibly affected.

In order to address these limitations, we decided to develop the Lethal Phenotypes Portal, an online web application, to showcase a curated catalogue of Mendelian genes with lethal phenotypes identified through the OMIM knowledge base. Here, we searched OMIM using a number of terms related to lethality, then collated and curated the resulting phenotypes. As previously noted, the definition and subsequent genes identified as lethal in humans can differ. Consequently, we categorised the entire set of OMIM disease associated genes into different lethality categories according to the earliest age of death reported using HPO age of death terms and definitions ^8,18^, from prenatal death to death in adulthood to genes with no reports of early death (non-lethal genes). In this manuscript, we characterise the genes across the different lethality categories, explore how this information combined with phenotypic similarity measures and gene family information could be used for novel gene discovery and examine the evidence on mouse viability. Additional visualisations available through the web tool allow to inspect how these categories correlate with other metrics on gene essentiality, including intolerance to LoF variation and cell gene effect scores.

## Results

### A comprehensive resource of genes with lethal phenotypes in humans Web application

The Lethal Phenotypes Portal is an online resource that provides users with a catalogue of human genes that are associated with documented lethal phenotypes in Mendelian disorders within OMIM. The web interface contains the full catalogue, allowing for queries and customised downloads, and a set of modules where genes in different lethality categories can be explored and compared with other sources of evidence on gene intolerance to LoF variation: constraint metrics, e.g., LOF Observed/Expected Upper-bound Fraction (LOEUF) from gnomAD and Selection against heterozygous (shet) scores inferred from 1 million genomes; gene effect scores from CRISPR cancer cell knockouts from DepMap and evidence on lethality from mouse knockout screens from the IMPC and phenotype annotations from MGI (see Methods). It shows a series of visualisations on the distribution of these metrics across different lethality categories (**Figure 3a**).

**Figure 3.**
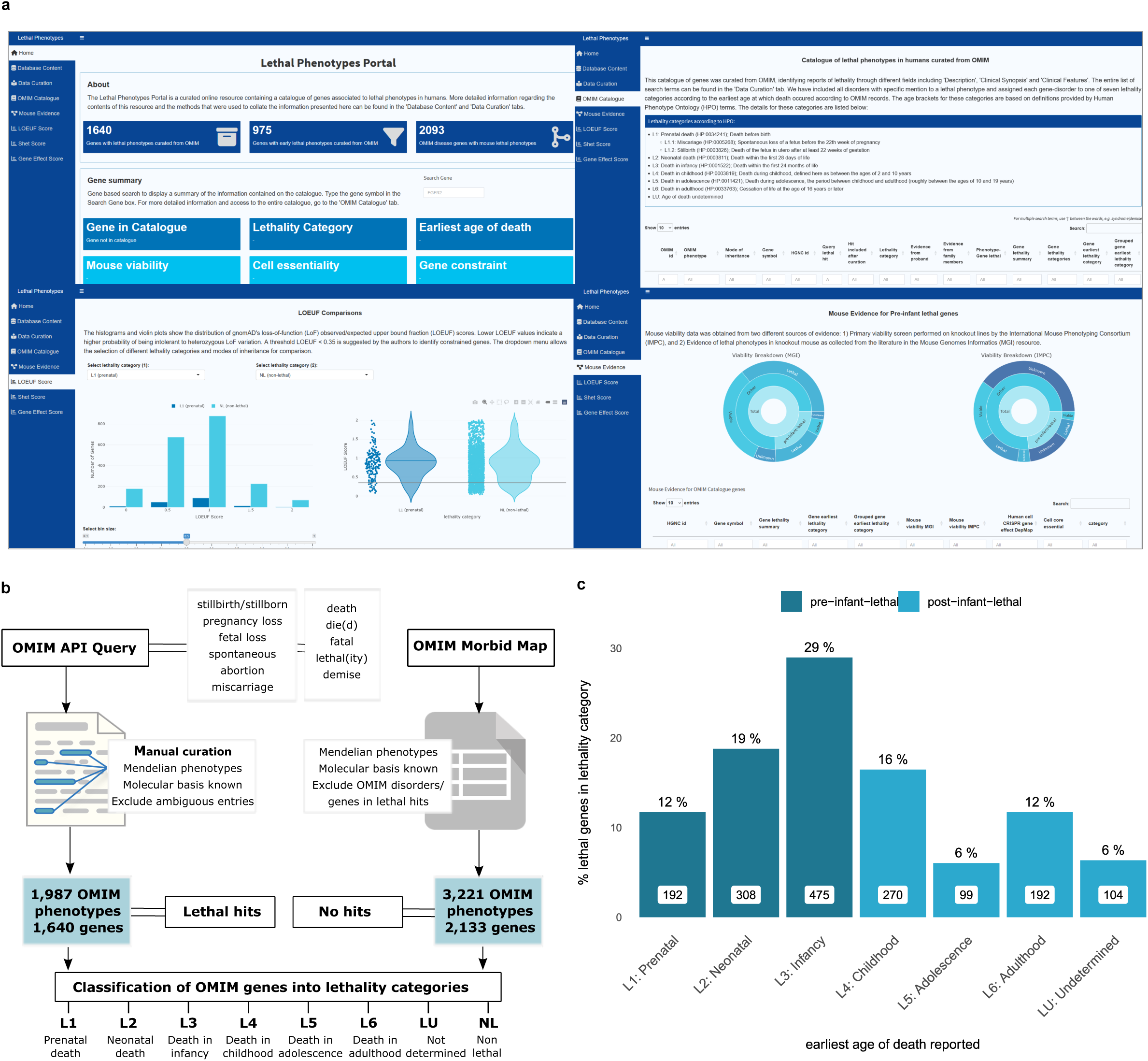
Catalogue query and curation strategy, data integration and web resource a) Web application. The resulting catalogue of lethal phentoypes is available to query and download through the following url: https://lethalphenotypes.research.its.qmul.ac.uk/. A set of annotations and visualisations allow the comparison between genes in different lethality categories in terms of intolerance to variation metrics, cell proliferation scores and mouse viability assessment **b) Query strategy** OMIM query strategy and curation pipeline to classify OMIM Mendelian phenotype associated genes into lethality categories **c) Distribution of OMIM lethal genes according to lethality categories** Bar plots represent the number (in white boxes) and percentage of genes in each lethality category with respect to all genes with records of early death. Genes with an associated earliest age of death in infancy are predominant among lethal genes.

The OMIM queries and curation constitute the main source of evidence on lethal phenotypes in humans captured in the resource. The outline for the query strategy and subsequent curation of the lethal phenotype hits is shown in **Figure 3b** and explained in detail in the Methods section and web application.

After manual curation and exclusion of ambiguous entries, we found that 57% (2,133/3,773) of genes associated with human single gene disorders catalogued in OMIM were not retrieved through the queries, suggesting no clinical records of lethality, 33% (1,239/3,773) are only associated to disorders with records of lethal phenotypes (as defined in Methods and **Figure 3b**), and 11% (401/3,773 are linked to both lethal and non-lethal phenotypes. With regards to lethality categories, 975 genes (59% of all lethal genes (1,640), 26% of disease genes) have records of prenatal, neonatal or infant death (pre-infant-lethal) as opposed to post-infant-lethal, where the earliest reported age of death ranges from childhood to adulthood. (**Figure 3b, 3c**, see Methods). The distribution of genes according to lethality categories are based on the earliest age of death reported.

### Characterisation of the set of lethal genes

Analysis of HPO annotations of the mode of inheritance revealed the genes linked to early death show a depletion of AD inheritance genes: 12% (118/975) of pre-infant-lethal genes are AD compared to 25% (165/665) of post-infant-lethal genes and 34% (719/2,133) of non-lethal genes (**Figure 4a**).

**Figure 4.**
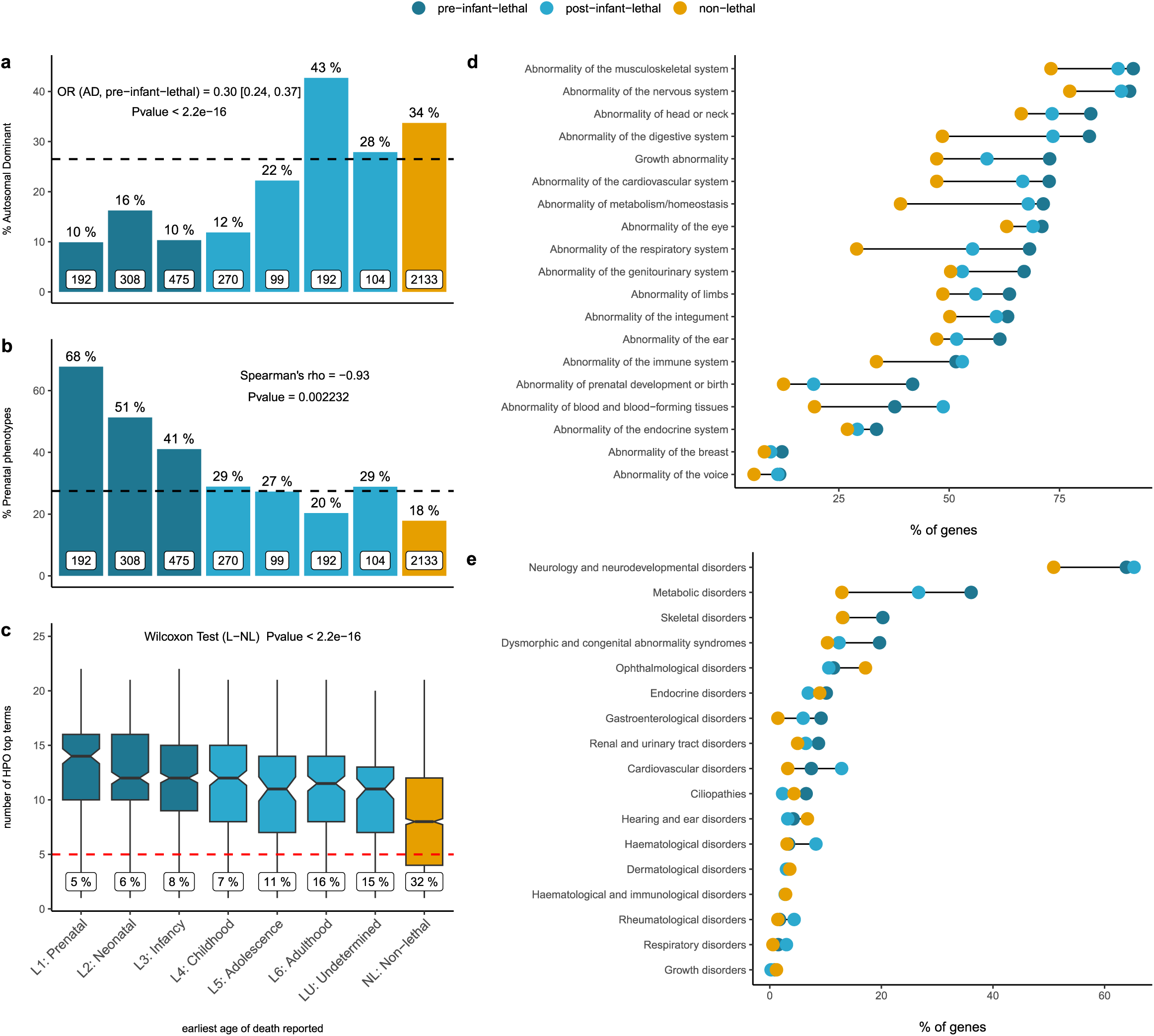
Human phenotype ontology and disease category analysis of genes in the catalogue a) Lethality categories and mode of inheritance. The set of pre-infant-lethal genes show a depletion of genes associated with an AD mode of inheritance. The bar plots represent the percentage of AD disease genes with respect to the total number of genes (white boxes) in each lethality category **b) Lethality categories and prenatal phenotypes** Bar plots represent the percentage of genes in each lethality category with abnormalities of prenatal development. There is a correlation between the earliest age of death reported and the presence of abnormal prenatal phenotypes **c) Lethality categories and abnormal phenotypes** The number of physiological systems affected is significantly higher among the lethal genes, implying more severe, multisystemic phenotypes (the numbers in the labels indicate the % of genes with HPO annotations mapping to <5 top HPO terms) **d) Lethality categories by affected systems** The percentage of genes in each category mapping to an specific physiological system is higher among the lethal genes for every single phenotype; **e) Lethality categories by disease group** The percentage of genes mapping to high level disease categories as per PanelApp (level 2 rare disease groups). OR: odds ratio; AD: autosomal dominant; L:lethal; NL: non-lethal; HPO: human phenotype ontology.

Exploring the prenatal phenotypes associated with the genes in the catalogue, i.e. those abnormal phenotypes under the ‘Abnormality of prenatal development or birth’ and ‘Intrauterine growth retardation’ parental terms, we observe a consistent trend across lethality categories. The earlier the age of death, the higher the likelihood of prenatal manifestation for a gene/disorder (**Figure 4b**).

The number of top-level HPO terms – phenotype terms that are direct descendants of the term ‘Phenotypic abnormality’ (HP:0000118) –, a proxy for the number of physiological systems affected, is significantly higher for the set of lethal genes compared to other disease associated genes, reflecting the multisystemic nature of these disorders and the presence of more severe clinical manifestations leading to premature death (**Figure 4c**). In accordance with this observation, the percentage of genes with an abnormal phenotype mapping to any of these individual systems is higher among those genes with records or early lethality compared to post-infant-lethal genes and non-lethal genes (**Figure 4d**). Similar patterns are observed when PanelApp disease classes are considered instead (**Figure 4e**). Consistent with the results reported using mouse viability data (**Figure 2d**), for ‘Ophthalmological disorders’ and ‘Hearing and ear disorders’ the percentage of genes is higher among the non-lethal category .

Interestingly, up to 26 % (250) of genes associated with disorders with pre-infant-lethality are also associated with other disorders with no records of lethal phenotypes. For 66 of these genes (26%), differences in allelic requirement could explain the differences in the severity of the phenotypes, since all the lethal forms are AR, and the associated non lethal disorders are AD (**Supplementary File 1**). Examples include *ACTL6B* or *ALG8* where, in addition, mouse mutants support this allelic model where the homozygous knockout shows embryonic lethality and the heterozygous knockout phenotypic abnormalities mimicking the phenotypes of the associated disorders (**Figure 5a**).

**Figure 5.**
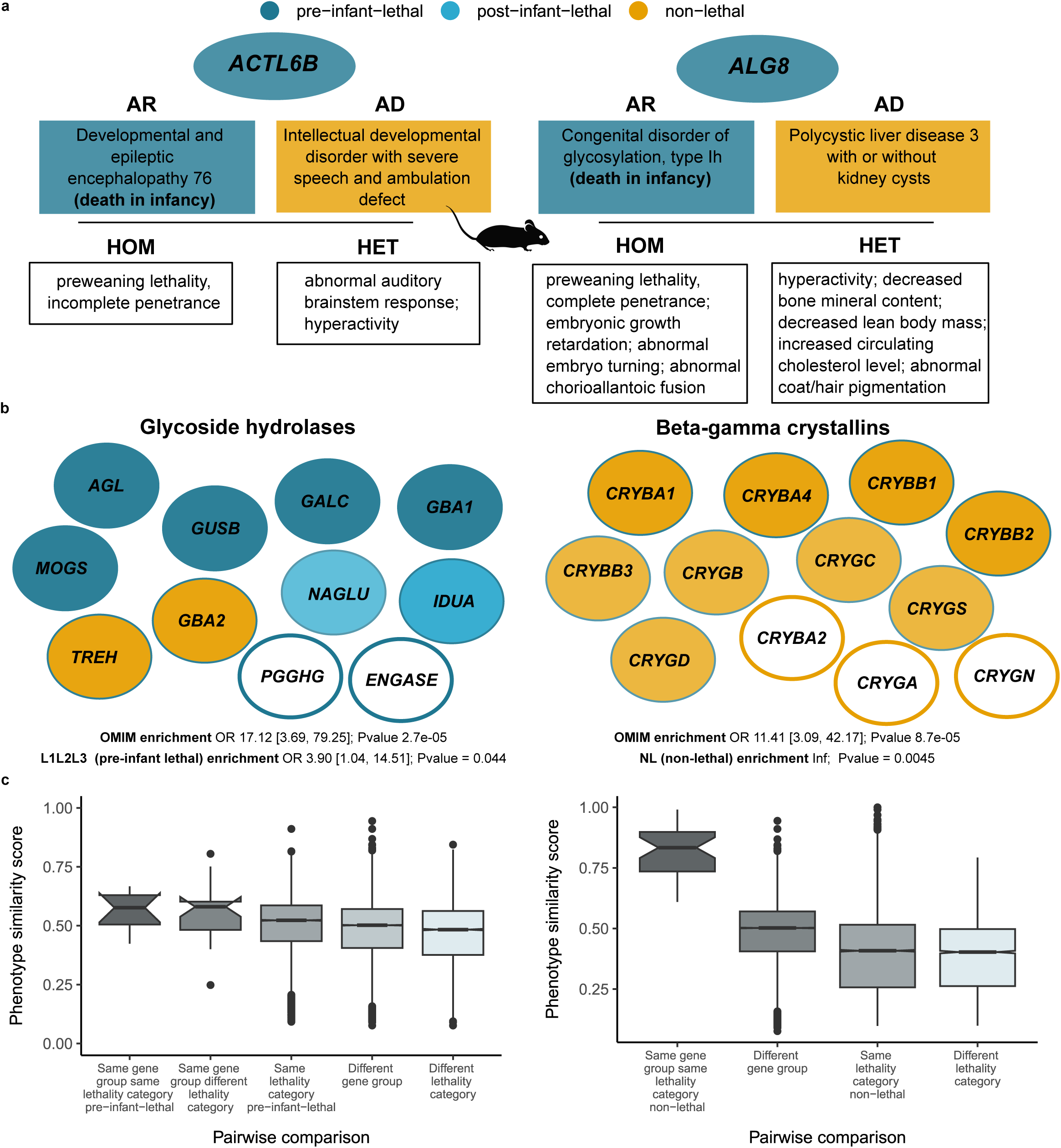
Gene group analysis of genes in the catalogue a) Genes associated to AR disorders with lethal phenotypes and AD disorders with no records of premature death. Mouse viability for the homozygous and heterozygous knockout is concordant with the phenotype observed in humans **b) Selected gene groups with potential novel candidate genes** Gene groups meeting the following criteria: 1) enriched for OMIM genes and 2) enriched for genes in lethality categories pre-infant-lethal (Glycoside hydrolases) and non-lethal (Beta-gamma crystallins) respectively. Those genes in white filled circles correspond to potential candidate genes to be associated to Mendelian conditions **c) Phenotypic similarity scores distribution of different pairwise comparisons for the two gene groups described in b)** Phenotype similarity scores between genes in the same gene group and lethality category compared to different subsets of genes belonging to different gene families and lethality categories. AR: autosomal recessive; AD: autosomal dominant; OR: odds ratio.

A further analysis of the causal variants would be needed to explore which other factors may help explain the spectrum of phenotypic severity, e.g. distinct location of variants (different protein domains); degree of functional impact (null alleles vs hypomorphic alleles); qualitative variation in functional impact, i.e. LoF vs gain-of-function (GoF) ^50^. It is worth noticing that GoF variation is more common in de novo/dominant disorders ^51^. The same variant can even be associated with different disorders or variations of a phenotypic spectrum, indicating other mechanisms need to be involved, e.g. gene-environment interactions, genomic imprinting, stochastic forces or genetic modifiers ^50,52^. Other factors that could explain variable penetrance comprise digenic/oligogenic inheritance, gene expression levels, age or gender ^53^. This once again reflects the challenges of trying to classify genes into binary categories, i.e. essential/lethal vs non-essential/non lethal and the need to build allelic-phenotypic series including prenatal phenotypes and age of death.

Finally, we investigated HGNC gene families/groups that are significantly enriched for both OMIM disease genes and one of the lethality categories (see details in Methods) and highlight two of them in **Figure 5b**. The ‘Glycoside hydrolases’ group was significantly enriched for pre-infant lethal genes and the group ‘Beta-gamma crystallins’ enriched for non-lethal genes. Genes in the same gene group/family not currently associated to Mendelian phenotypes are suggested as candidates: *PGGHG* and *ENGASE,* and *CRYBA2* (currently with a phenotype association reported as provisional in OMIM), *CRYGA*, and *CRYGN* respectively. The complete gene list is available in **Supplementary File 2**. This finding is supported by the analysis of phenotypic similarity scores for the genes in these two groups where genes within the same gene group and with the same pre-infant lethality category showed higher similarities compared to other genes in different groups and lethality categories (**Figure 5c**). Phenotypic-driven, variant prioritisation algorithms, e.g. Exomiser ^54^, are already used to identify diagnostic variants and could be expanded to identify variants in potentially novel disease genes where the gene belongs to the same gene group and lethality category as a known disease gene with associated phenotypes similar to the patient being investigated.

### How well does essentiality correlate between organisms?

Using the set of genes associated with prenatal and neonatal lethality in humans and combining it with viability data from the mouse orthologs, we can look at the overlap between the sets of lethal genes in the two species. Out of 438 pre-infant lethal genes in humans with IMPC mouse orthologue data on viability, 322 are also lethal in the mouse, while 116 genes are mouse viable, which implies a discrepancy of 26% between the two organisms in terms of gene essentiality.

This percentage is slightly lower if we include only pre- and perinatal death (22%). By contrast, we find AR disease genes, where no records of premature death were captured and the corresponding mouse ortholog is lethal (256/586, 44%). Some of the hypothesised reasons behind these discrepancies are highlighted in **Figure 6**. These range from differences in the type of genetic variants and mechanisms (LoF versus non-LoF) to variable transcriptional and functional compensation mechanisms. For the disease genes in the other two categories (post-infant-lethal and AD disease genes with no records of lethal phenotypes) the differences in lethality could be more easily explained, i.e. deficit of homozygous variants leading to embryonic lethality in humans.

**Figure 6.**
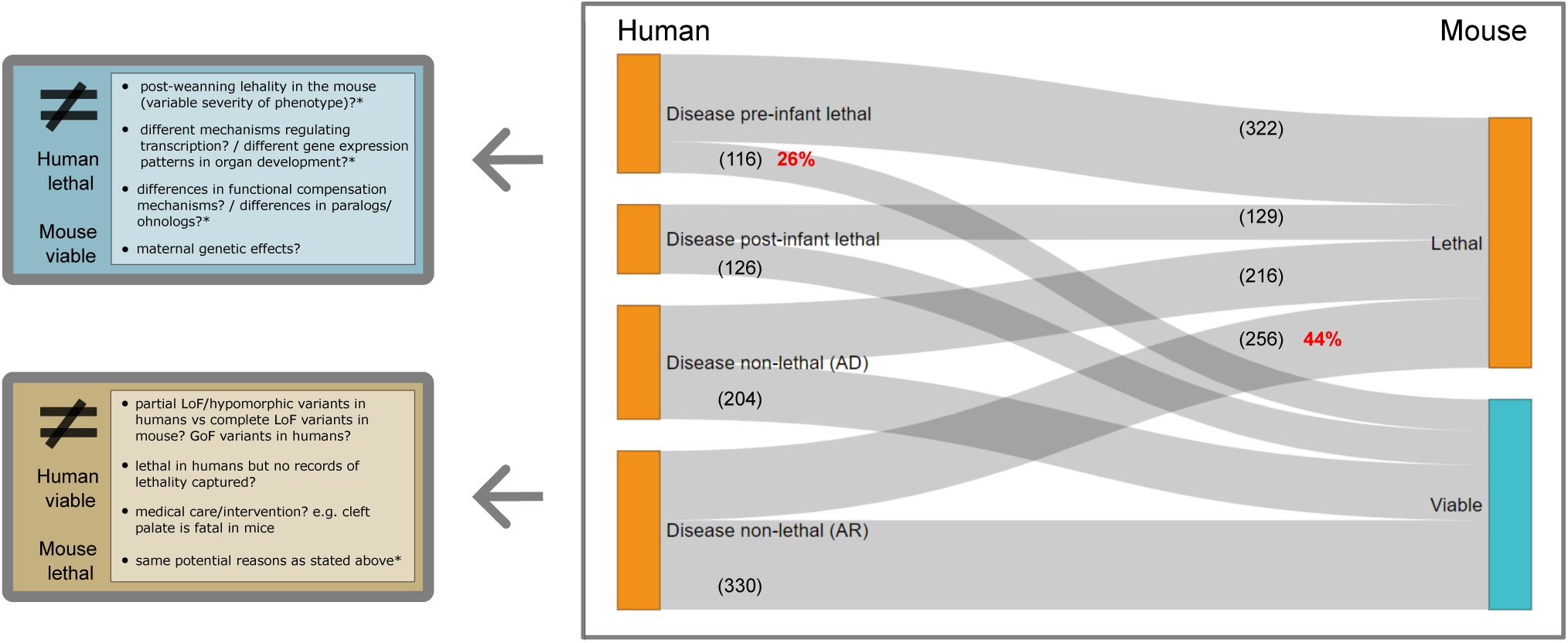
Evidence of lethality in human and mouse and potential reasons for discrepancies. Human disease genes in the catalogue with a one-to-one mouse ortholog that has undergone the IMPC primary viability assessment (DR 20.1). Discrepancies in viability between the two organisms are highlighted, together with multiple hypothesis that could explain these differences for the two most extreme scenarios: 1) pre-infant lethal phenotypes in humans and pre-weaning viability in mouse, and 2) AR disease genes with no records of premature death in humans and pre-weaning lethal phenotypes in the mouse (complete or incomplete penetrance, i.e. lethal + subviable). AD: autosomal dominant; AR: autosomal recessive; LoF: loss-of-function; GoF: gain-of-function.

Of the total number of genes with a lethal phenotype in knockout mice and a one-to-one human orthologue (IMPC and MGI combined, 5,064), up to 54% have no phenotype associations reported in humans to date according to OMIM. Given the strong and consistent evidence of the association of lethal genes in the mouse and disease genes in humans, these 2,721 genes represent a substantial source of potential candidates for Mendelian disorders, including prenatal conditions ^21^. This information, combined with other sources of gene essentiality as displayed in the web application can assist the prioritisation of variants in novel genes.

## Discussion

### Relevance of the resource

Essential genes and Mendelian (lethal) genes are not two independent concepts. The experimental evidence we have on essential genes comes mainly from cell proliferation assays and model organism viability studies. The current sources of human lethal phenotypes consist of single-gene disorders repositories, since the disruption of a gene function leading to embryonic/prenatal lethality can be interpreted as the most severe manifestation of these disorders. However, these phenotypes, and their associated genes, are not comprehensively captured in current databases. Here we queried and curated lethal phenotypes described in the OMIM catalogue to categorise human disease genes, using HPO terms, according to the earliest reported age of death. We integrated this data with metrics on gene constraint inferred from human population sequencing data, cell and mouse essentiality status, and provide a number of user-interactive visual and analytical features. In making this resource openly available to the public, we hope that this application will be used as a tool to aid clinical geneticists in diagnosing early lethal conditions, allowing better informed pre- and perinatal counselling and family planning. Additionally, it will assist researchers investigating what makes these genes so essential for human development, and at what stage.

We also describe a characterisation of the set of lethal and non-lethal genes in humans, highlighting potential strategies for novel Mendelian gene discovery. It is unlikely that we have identified most of the genes associated with rare disorders ^24^, let alone all the monogenic forms of embryonic loss (before a pregnancy is recognised) and fetal death, that once again, may be considered an extreme manifestation of some Mendelian conditions. Information on lethality category, gene group annotations, and phenotypic similarity between undiagnosed patients and known disorders or among patients could be integrated for this purpose. Strategies for variant prioritisation leveraging information from other members of the gene group have previously been successfully implemented ^55^. When assessing the significance of potentially pathogenic variants in unknown disease genes, evidence of lethality in mice in combination with intolerance to variation metrics have independently been used to prioritise candidate variants in potential novel genes ^5,37,49^.

### What have we learned about lethal phenotypes in humans?

Analysis of the phenotype annotations of the genes, categorised by earliest age of death for their associated disorders, revealed several correlations. First, the proportion of AD disease genes is significantly lower for pre-infant lethal genes. Second, the number of physiological systems affected is significantly higher when we compare lethal vs non-lethal genes. Third, there is a significant correlation between lethality category and presence of prenatal abnormalities. In terms of disease categories, metabolic, dysmorphic and congenital abnormality syndromes and skeletal disorders are more frequent among pre-infant lethal genes compared to post-infant lethal and non-lethal genes. The classification used for this analysis based on PanelApp disease categories presents potential bias due to how broad some groups are compared to others, as well as potential overlaps. It is also important to mention that, prenatally, organ system anomalies is what can be most effectively assessed and compared, and these prenatal phenotypes could be related to different categories, e.g. fetal hydrops, a unique severe often lethal prenatal phenotype can be associated to cardiovascular, immunological, or the haematological category. More granular associations can also be found when we explore the exact embryonic stage at which the mouse embryo dies ^30^.

### Comparison between mouse and human

Comparing the set of essential genes in different species is a particularly challenging task. Even within the same species, several factors may affect the essentiality assessment: the specific developmental stage at which viability is evaluated, the exact measure of viability (survival vs fitness), the assessment approach (inference, e.g. absence of biallelic complete LoF mutations in population sequence data versus observation, e.g. molecular autopsy), environmental conditions and genetic background ^5,56^.

The mouse is the most extensively used model organism in the study of human disease, particularly in the context of single-gene disorders, allowing us to explore how genetic variants impact the phenotype ^57,58^. However, the ability of mouse models to capture human phenotypes is not without limitations ^59^. When performing comparisons between mouse and human lethal genes, several considerations need to be taken into account: 1) RNA expression profiles, including differences in developmental gene expression may reflect physiological differences between these two organisms ^60,61^. This is supported by evidence of evolutionary divergence in regulatory networks contributing to phenotypic differences ^62,63^; 2) molecular function and biological processes are likely to remain constant between different species while physiological relevance may differ ^64^; 3) based on that, we may want to consider essential functions instead of essential genes, and approaches based on projection over functional modules, e.g. pathways or networks, have already been implemented ^56^; 4) even within the same species, we may observe variability due to genetic background, e.g. differences in lethality were found for up to 10% of mouse knockouts with data on viability ^5^. Additionally, lethality might show incomplete penetrance even with the same mutation and genetic background . Taking all these factors into account, a 75% overlap between pre-infant lethal and mouse lethal genes, together with the fact that those disease categories where prenatal and neonatal lethal phenotypes in humans are more frequently reported show a higher percentage of genes with an ortholog mouse knockout that is also lethal at embryonic and pre-weaning developmental stages, are indicative of concordant cross-species phenotypic effects with certain degree of variability. This percentage of genes with discordant phenotypes between the two species is consistent with previous findings ^64^. Overall, while differences in essentiality between mouse and human are undoubtedly expected to some extent, to date, the set of mouse lethal genes remains our most valuable source of information on genes essential for mammalian development. The evidence is strong and consistent regarding the association of lethal genes in the mouse and disease genes in humans ^1,5,65^. Consequently, the set of mouse lethal genes with no existing human evidence constitutes a powerful source of potential candidates for Mendelian disorders, including those with prenatal/neonatal lethal phenotypes.

### Challenges and limitations

One of the main limitations is the potential for false negatives when determining the lethality category associated with each human disease gene. First, the queries may have failed to detect all the records of death described in OMIM. Second, and a more likely factor leading to misclassification of genes, is that the curation is limited to the information captured in OMIM, and does not include the original source. A gene classified as non-lethal indicates that we were not able to retrieve any record of lethality in OMIM, and thus the non-lethal phenotype category may be overestimated. Third, there is a manual curation component that is prone to human error and/or biased interpretation (e.g., cause and age of death is ambiguous; death may refer to siblings or other family members with variable evidence of being affected by the same disorder of the proband).

Allele type is an important caveat. Clearly we do not have a full view of genome-wide nullizygosity in humans, only inferences from constraint and heterogenous reports of disease-associated lethality, the latter of which is likely to be impacted by the nature of the reported alleles and the specific mechanisms. It is worth noting that for some of the gene-disease associations captured in the catalogue, the mechanisms may not be necessary one of LoF, e.g. GoF mutations also lead to lethal phenotypes ^66,67^. Similarly, within the set of AD disease associated genes we may find both de novo and inherited monoallelic variants. Lastly, recent molecular autopsies studies are not necessarily captured in OMIM, since there is a gap between a novel gene disease association being published and captured by this repository. In the same manner, brief reports of expansion of phenotypes to include prenatal lethality may not necessarily be reflected.

### Further plans

The increasing number of cases undergoing prenatal and neonatal sequencing and molecular autopsies will reveal novel Mendelian genes as well as expansion of the phenotypic spectrum for other known disease genes. Complementing this resource with a literature review of these studies might add a set of potential candidate genes to be associated to prenatal and neonatal lethal phenotypes where predicted pathogenic variants in novel genes are identified ^37,48^. For those cases where early death constitutes a novel phenotype in a known disease gene, we will focus on generating allelic-phenotypic series and establish correlations between variants/gene/protein features and the clinical manifestations observed in patients.

Other categories of essential genes could be incorporated in this catalogue. A comprehensive resource of genes and variants associated with infertility in different model organisms and humans has recently been published ^68^. Infertility is an emerging public health issue as 10-20% of couples are infertile worldwide and global fertility rates are falling ^69^. Approximately 30-40% of infertility cases are of unknown aetiology ^70,71^. The contribution of essential/lethal genes to clinical infertility due to recurrent embryonic loss at the very earliest stages before a pregnancy is recognised is currently underappreciated, but we would expect them to explain a proportion of these cases, e.g. genes affecting zygotic and early cleavage stage survival ^72,73^. Records of lethality for some of these genes included in the catalogue are often heterogenous and/or ambiguous and should be interpreted with caution.

Similarly, it would be useful to create specific categories for maternal effect genes (MEG) associated with clinical infertility due to early embryonic loss as well as infertility due to abnormal oocyte development. Miscarriage constitutes another recognised phenotype for some MEGs, in particular due to recurrent molar pregnancy ^74^. An additional category could include those genes where LoF may be associated with other phenotypes linked to reduced reproductive success ^75^. The information curated for this study is currently in the process of being incorporated into the HPO resource. Overall, this catalogue represents one more step towards eventually categorising all human genes across the full spectrum of intolerance to variation: from genes with pathogenic variants leading to early embryonic death to rare, genuine, homozygous LoF variants found in healthy adult individuals.

## Methods

### Data collection

#### OMIM Data

Disease-gene associations were data mined by using the OMIM API ^17^ [https://www.omim.org; Data last accessed 24/11/23] to search terms linked to lethality. Manual curation of all the hits was performed and a series of inclusion and exclusion criteria were applied to discard ambiguous reports of lethality. The list of terms and summary of the OMIM data curation process is illustrated in **Figure 1**. The initial queries date back from May 2020, subsequent queries and curation of hits was performed, including the inspection of 10% of previous curated entries with potential updates. This implied the reclassifications of some lethality categories, mainly between L1 to L3 and L6 to LU labels. The OMIM Morbid Map was pruned to exclude provisional gene-phenotype relationships, non-diseases and drug response phenotypes. Disorders with somatic, multifactorial and digenic modes of inheritance were also excluded. As a result, the gene-phenotype associations included in the catalogue are limited to Mendelian phenotypes with molecular basis known and where the gene is classified as protein coding according to HGNC ^76^.

The initial hits that were not included after manual curation were labelled as ambiguous entries. Each unique disease-gene association was assigned to a ‘lethality category’ based on the earliest time point in which lethality had been documented to occur, with categories grouped by age ranges defined by the HPO age of death categories ^18^. The exact definitions of each lethality category set and more details on the entirety of the OMIM curation can be found in the web application. Additional information clarifying whether the evidence of a lethal phenotype comes from a proband or a family member, e.g. history of miscarriages in the family, is also included.

#### Gene Properties and Essentiality Annotations

HGNC ids and information on gene groups ^76^ [https://www.genenames.org/; Data accessed 19/12/23], gene-disease associations according to OMIM and their associated mode of inheritance(s) and abnormal phenotypes according to HPO annotations [https://hpo.jax.org/app/data/annotations; Data accessed 19/12/23] were retrieved for all human protein coding genes. Information on disease categories was retrieved from Genomics England PanelApp API ^27^ [https://panelapp.genomicsengland.co.uk/api/v1/genes/; Data accessed 19/12/23].

Several additional metrics and properties of essentiality for each gene were collected: Intolerance to LoF variation gene-level metrics from gnomAD v4 ^6^ [https://gnomad.broadinstitute.org/], Selection coefficients on fitness from RGC-ME ^14^, Gene viability data from mouse orthologs from the IMPC web portal ^22^ [https://www.mousephenotype.org/; mouse viability assessment, DR 20.1, Data accessed 15/12/23] and the MGI database ^7^ querying lethal phenotypes from Dickinson et al. ^1^, [https://www.informatics.jax.org/; Data accessed 15/12/23] and Human cell line proliferation scores according to the Cancer Dependency Map Portal’s Project Achilles ^9^ [DepMap 23Q4 CRISPR Gene Effect score, https://depmap.org/portal/ ; Data accessed 15/12/23].

Phenotypic similarity scores between Mendelian disorders were computed using PhenoDigm algorithm ^23^ and HPO annotations ^18^ [https://hpo.jax.org ; Data accessed 04/07/23].

### Database organisation / Web application

The gene annotations were ultimately organised into two main files within the Lethal Phenotypes Portal: OMIM curation and gene annotations. The web application was built using R programming language (v4.3.1) ^77^, *‘shiny’*(v.1.7.5) ^78^ and *‘shinydashboard’* (v0.7.2) ^79^, which allows for the relevant information contained in the catalogue to be presented in a dashboard format. The *‘plotly’* (v4.10.2) R package ^80^ was used to create visualizations of interactive plots and *‘DT’* (v0.28) R package ^81^ as an interface to the DataTables JavaScript library to display the resulting datasets. Other packages used include *‘dplyr’* (v1.1.2) ^82^ and *‘stringr’* (v1.5.0) ^83^.

### Data analysis

The figures in the manuscript were created using *‘ggplot2’* ^84^ and *‘networkD3’* ^85^ R packages. All the statistical analyses including Odds Ratios and Fisher test, correlation coefficients, Mann-Whitney test were performed in R ^77^.

Gene family enrichment analysis: 1,053 out of 1,509 gene groups provided by HGNC include at least one gene present in the catalogue curated from OMIM (subset of genes associated with Mendelian phenotypes with molecular basis known). For each one these gene groups, the proportion of genes in the catalogue was computed, and for those genes in the catalogue, the proportion of genes in each ‘merged’ lethality category: pre-infant lethal, post-infant lethal and non-lethal. Odds Ratio, CI and Fisher test p-values were computed for each group to identify gene groups enriched in OMIM genes and any lethality category. Uncorrected p-values are shown.

Phenotypic similarity analysis: For each OMIM disorder, the associated HPO phenotypes are retrieved and the phenotypic similarity for all disease-disease pairwise combinations is computed. Each disorder is then mapped to its associated gene/s to obtain gene-gene scores. The distribution of phenotype similarity scores for genes in a given gene group can be compared with similarity scores for different subsets: catalogue genes belonging to the same gene family and same lethality category, catalogue genes belonging to the same gene family and different lethality category, catalogue genes belonging to the same lethality category (regardless of gene group), catalogue genes belonging to the same gene group (regardless of lethality category), catalogue genes belonging to different gene group, catalogue genes belonging to different lethality category.

The web interface is available for data exploration at https://lethalphenotypes.research.its.qmul.ac.uk

Supplementary Files accompanying this manuscript can be found in the following repository: https://zenodo.org/records/10419108

## Acknowledgments

This research utilised Queen Mary’s Apocrita HPC facility, supported by QMUL Research-IT: http://doi.org/10.5281/zenodo.438045. We are grateful to present and past members of the QMUL ITS Research team (Tom Bradford, Giles Greenway, Iain Stenson, Iain Barrass) for their help with the development and deployment of the Shiny app.

## Authors’ contributions

P.C. contributed to conceptualisation, data analysis, presentation and interpretation of the results and writing the manuscript. S.L. contributed to data analysis and results interpretation, reviewing and editing the manuscript. I.B.V.d.V., D.Z., M.D., P.N.R. contributed to reviewing and editing the manuscript. G.B. contributed to data analysis and reviewing the manuscript. S.A.M. contributed to data generation, reviewing and editing the manuscript. D.S. contributed to conceptualisation, data analysis, interpretation of the results and reviewing and editing the manuscript. D.S., S.A.M. are PIs of the key programmes who contributed to the management and execution of the work

